# INSIGHTFUL: Insight Generation through Clinical Annotation, Analysis, and Modeling of Suicide-Related Factors towards Understanding and Lifesaving

**DOI:** 10.1101/2025.01.13.25320491

**Authors:** Zehan Li, Rodrigo M Vieira, Sunyang Fu, Yan Hu, Wanjing Wang, Andrew Wen, Scott Lane, Salih Selek, Lokesh Shahani, Hua Xu, Jair C Soares, Hongfang Liu, Ming Huang

**Affiliations:** MacWilliam School of Biomedical Informatics, The University of Texas Health Science Center at Houston, TX, USA; Department of Psychiatry & Behavioral Sciences, McGovern Medical School, The University of Texas Health Science at Houston, Houston, TX, USA; Section of Biomedical Informatics and Data Science, School of Medicine, Yale University, New Haven, CT, USA

## Abstract

**Objective:** Suicide is a critical medical and public health challenge, particularly among individuals with mental illnesses in safety-net hospitals. To uncover insights about suicidality embedded in unstructured clinical notes, we propose to annotate, analyze, and model a corpus for suicidality understanding and lifesaving.

**Methods:** A multidisciplinary panel developed an annotation guideline to capture four key suicide-related factors: Suicidal Ideation (SI), Suicide Attempt (SA), Exposure to Suicide (ES), and Non-Suicidal Self-Injury (NSSI). We created an annotated corpus of 500 notes through a clinically validated annotation process and performed cohort analysis to characterize demographic and suicidal distributions. A large language model was deployed for automatic classification.

**Results:** The annotated corpus was created with a Cohen’s Kappa of 0.95 and further de-identified for data sharing. Most notes (79.4%) contained one (34.4%) or more (45%) suicide-related labels, with SI and SA co-occurrence as the most frequent combination (35.6%), which demonstrates significant overlap. The cohort was characterized with a mean age of 33.4, 51.7% male, and 75.8% singles. Prevalent stressors included unemployment (24.2%), homelessness (12.0%), limited healthcare access (5.4%), and legal challenges (5.0%). We identified four key insights to improve documenting suicidality, including implicitness, confliction, ambiguity, and definition coverage incompleteness. The baseline model achieved a micro-averaged F1 score of 0.70, demonstrating satisfying performance in multi-label classification.

**Conclusion:** The near-perfect inter-annotator agreement underscores the proposed annotation process and data quality. Cohort analysis highlights the distribution and documentation insights of suicidality. Data modeling demonstrates the potential of insight generation via AI-powered methods for mining large-scale clinical notes.

## Introduction

Suicide is a pressing and complex medical and public health challenge, especially among individuals with mental illnesses (1–3). Highlighting the magnitude of this crisis, Centers for Disease Control and Prevention (CDC) reported that 48,183 people died by suicide in the United States, while 12.3 million adults seriously thought about suicide, 3.5 million developed a plan, and 1.7 million attempted suicides in 2021 (4). Suicidality often involve multiple stages, from ideation to planning and attempting suicide, and is influenced by multiple factors such as age, gender, family history, socioeconomic status, and broader socio-political circumstances, reflecting the challenges of suicidality management in clinical care (5–7).

The widespread applications of EHR systems allows the enhancement of the identification and intervention of suicidality in psychiatric care settings, particularly within safety-net hospitals (2, 8). Although traditional methods often relied on self-reported surveys or manual chart reviews of clinical diagnoses captured in administrative data, EHRs present unprecedented opportunities for tracking standardized and coded suicide-related behaviors (9, 10). However, systematic reviews show that suicidal events and related factors are often under-coded in structured EHR data, resulting in low sensitivity when identifying and analyzing suicide or suicidal ideation (11, 12). To overcome the limitations of structured EHR data, the rich narratives contained within clinical documentation provide an invaluable alternative resource for suicide management. Clinical notes, such as initial psychiatric evaluation (IPE), capture detailed patient mental health status, life stressors, and behaviors, offering a systematic view of suicidality (13, 14). The unstructured clinical narratives often require manual content analysis of a small sample of clinical notes for a deep understanding of suicidality (15). The manual analysis procedure includes the panel construction, annotation guideline development, corpus annotation, and content analysis. However, the manual content analysis is time-consuming and costly and is very challenging for processing large-scale clinical documents for observational studies. With Artificial Intelligence (AI) technology, Natural Language Processing (NLP) provides a feasible and effective solution for processing a large collection of clinical notes. By leveraging NLP methods to extract suicidal events and related factors from psychiatric evaluation notes, there is a significant opportunity to address the under-reporting issues found in structured billing data and enhance risk assessment efforts in clinical practice (16–18).

One caveat of NLP is the need for high-quality data derived from expertly annotated corpora. The effectiveness of NLP tools heavily relies on the accuracy and relevance of the training data, making the development of comprehensive, domain-specific annotated datasets essential for reliable outcomes (19–21). Most recent work in applying NLP for suicide detection, such as rule-based and deep learning methods, has focused on social media data, which, while valuable for studying general patterns of self-reported suicidal events and related factors, lacks the rigor and clinical reasoning required for healthcare settings (22–24). The lack of clinically validated annotation guidelines and an abundance of annotated clinical text represent significant bottlenecks in the field. Without these resources, NLP models risk misinterpretation or oversimplification of nuanced clinical language, which can lead to reduced performance and reliability when applied to real-world medical cases (21, 25). Additionally, the variability in clinical narratives across different healthcare systems and practitioner styles further complicates the creation of standardized annotation protocols (26, 27). Addressing this challenge requires collaborative efforts between NLP experts, clinicians, and mental health researchers to develop robust, clinically meaningful annotation standards that accurately reflect the complexity of patient documentation. The creation of such high-quality annotated datasets would empower NLP models to better extract and interpret crucial details from psychiatric notes, ultimately enhancing the detection and understanding of suicide risk within clinical care environments.

To further our understanding of suicidal events and related factors documented in clinical notes within the context of psychiatric safety-net hospitals, we propose to create a golden standard corpus of IPE notes on suicidality, perform a manual content analysis of this sampled dataset, and deploy NLP algorithms for text analysis automation. Specifically, we formed a panel of experts in psychiatry and biomedical informatics. This panel developed a clinically sound annotation guideline and constructed an annotated corpus using the IPE notes from a local safety-net psychiatric hospital in Harris County, Texas. We performed cohort and content analysis on 500 notes to characterize the demographics and label distributions and documentation patterns. We further developed and evaluated a pretrained language model as a baseline which enables data mining of a large collection of clinical documents.

## Methods

As illustrated in **Figure 1**, we curated a sample of 500 Initial Psychiatric Evaluation (IPE) notes from a longitudinal electronic health record (EHR) database. An annotation guideline was developed through an iterative process, incorporating clinician expertise, clinical knowledge, and sample notes. The corpus was then annotated for the presence or absence of four suicide events and related factors. We conducted a quantitative analysis of the study cohort and annotated corpus, along with a qualitative content analysis to generate insights for clinical practice and biomedical informatics research. Additionally, we evaluated the performance of AI-powered classification to assess the feasibility of automatically detecting suicide-related events in clinical notes using a multi-label classification approach. The following sections will provide a detailed discussion of each step.

**Figure 1.**
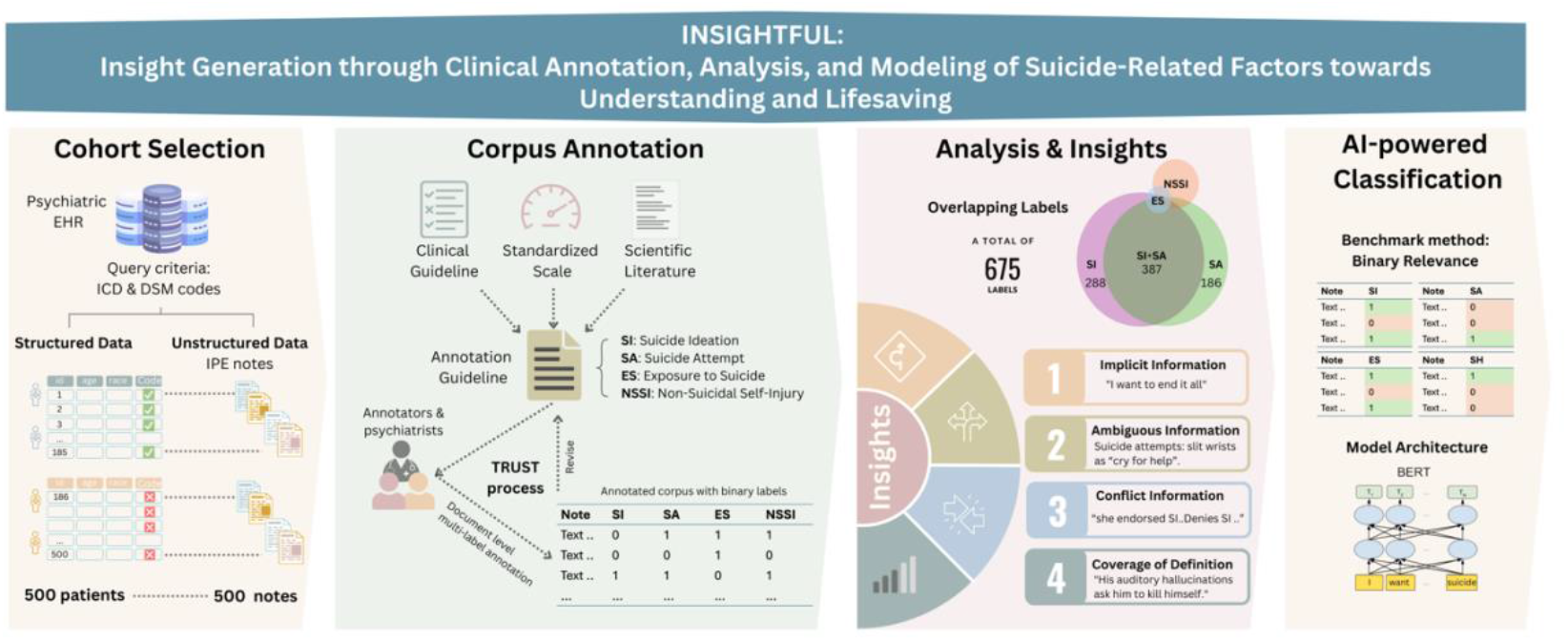
Graphical abstract detailing study workflow from cohort selection, corpus annotation, cohort analysis and insight generation, and classification automation

### Data collection

We accessed a comprehensive repository of EHR data from the Harris County Psychiatric Center (HCPC) at the University of Texas Health Sciences Center at Houston. This longitudinal EHR database spans from 2001 to 2021 and includes millions of clinical notes from 118,421 patients. We constructed a study cohort of 500 patients and a corpus consisting of 500 IPE notes for sequential analysis. Of these 500 records, only 185 were identified with structured fields indicating any suicide-related tendencies. The 315 records were randomly selected from the remaining patient records without structured documentation.

### Annotation Guideline Development

An expert panel consisting of psychiatrists and mental health researchers provided oversight for the development of annotation guidelines and data annotation process. The initial annotation guidelines were developed based on three authoritative sources: clinical guidelines, standardized scales, and scientific literature. Specifically, the definitions of suicidal ideation (SI), suicide attempt (SA), and non-suicidal self-injury (NSSI) were derived from Wolters Kluwer’s UpToDate and Columbia Classification Algorithm of Suicide Assessment (C-CASA) (28, 29). Although exposure to suicide (ES) as a suicide-related factor is not defined in clinical guidelines, we adopted definitions from a meta-analysis (30). We developed five general rules after conducting a manual review of 50 IPE notes (See Annotation Guideline in Supplemental material for details). For example, the guideline prioritizes physician-reported evidence over patient self-reports in cases of conflicting information, addressing the common clinical challenge where patients may minimize or deny their suicidal tendencies despite clear clinical evidence to the contrary. Label-specific rules and working definitions were further refined after an additional 150 notes were reviewed. For example, when SI arises not from the conscious mind but because of psychiatric conditions, such as auditory hallucinations commanding the patient to end life, these instances are classified as positive SI cases.

### Data Annotation

Following the Clinical Text Retrieval and Use for Scientific rigor and Transparent (TRUST) process, two annotators applied the annotation guidelines to the 500 IPE notes, including the 200 notes used for guideline development (See **Figure 2**) (31). Each label category was classified using a binary code, where ‘1’ indicates a positive mention and ‘0’ indicates a negative mention. This guideline was subsequently further refined through an iterative annotation and adjudication process involving the two annotators and three board-certified psychiatrists. We assess the quality of annotation using Inter Annotator Agreement (IAA) at two stages: (1) at the end of the guideline development with 200 notes and (2) the initial completion of annotating all 500 notes. The remaining disagreements were adjudicated and resolved through discussion with a panel of 4 board-certified psychiatrists. This iterative approach ensures clinical accuracy and comprehensiveness of the guideline development and corpus annotation.

**Figure 2.**
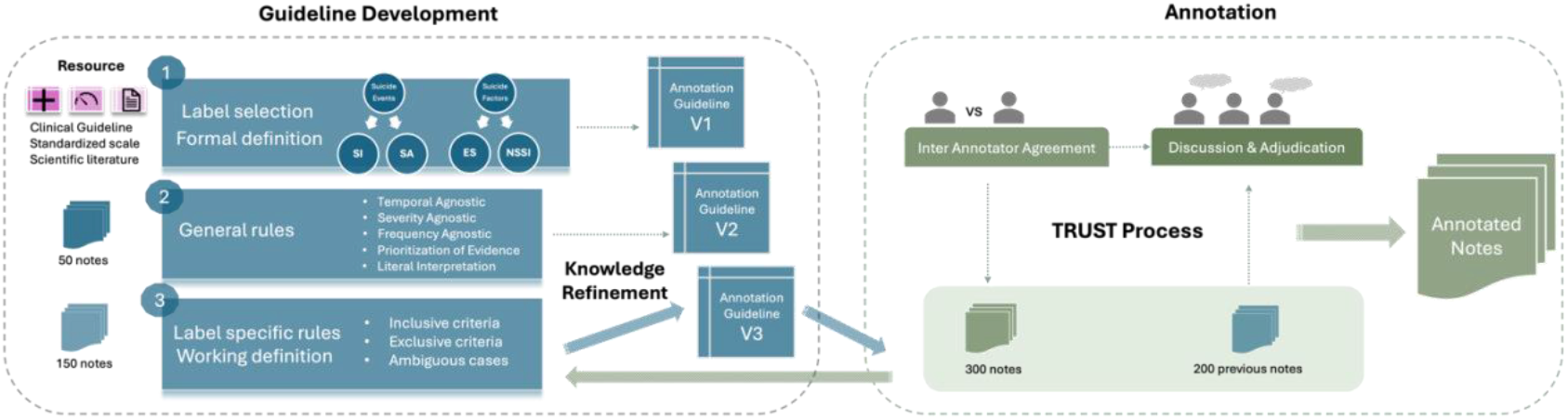
Annotation guideline development and corpus annotation procedure

### Data Analysis

We started with a demographic analysis of the study cohort with respect to the four suicide-related labels and patients including non-suicidal (NS) patients via descriptive statistics. The demographics includes age, gender, marital status, and race or ethnicity, together with social stressors extracted as diagnosis codes in the structured EHR. We also performed corpus analysis by calculating inter annotator agreement (IAA) and label distribution and content analysis to examine suicidality documentation. Refer to the **Supplement** for a detailed selection for social stressors.

### Data Modeling

We developed and evaluated an AI-powered multi-label classification approach to automatically identify coexisting suicidal events and related factors from IPE notes, by utilizing pretrained language models based on Bidirectional Encoder Representations from Transformers (BERT) (32). The multi-label classification was implemented via a binary relevance method, also known as One-vs-All, where each suicidal event was treated as a separate binary classification task (33). The pretrained language models were fine-tuned with a training set of 400 IPE notes and evaluated with a testing set of 100 remining notes. The training and evaluation were performed using 5-fold cross-validation with 3 repetitions. We calculated accuracy and micro-averaged precision, recall, and F1 scores for model performance evaluation.

### Data De-identification and Sharing

We adhered to NIH Safe Harbor guidelines for de-identification of personal identifiers in the (34). All 500 IPE notes were de-identified and made available for access under the data use agreement of UTHealth.

## Results

### Cohort Demographics

Within the study cohort of 500 patients, patient ages ranged from 6 to 82 years, with a mean age of 33.42. Patients with NSSI had a lower mean age of 26.90 years, highlighting a concerning trend toward younger demographics. Gender distribution was relatively balanced, with a slight male predominance (53.4%) over females (44.6%). Most patients were White (59.60%) or Black (30.60%), with smaller proportions of Hispanic (5.0%) and Asian (1.40%). A majority (75.80%) were single, suggesting potential social isolation linked to psychiatric conditions and suicidal tendencies. Economic and social factors were prominent, with unemployment (24.20%), homelessness (12.00%), limited healthcare access (5.40%), and legal challenges (5.00%) affecting many patients. The cohort’s average length of stay (LoS) was 9.52 days, reflecting the high-acuity, safety-net nature of HCPC’s operations.

**Table 1.**
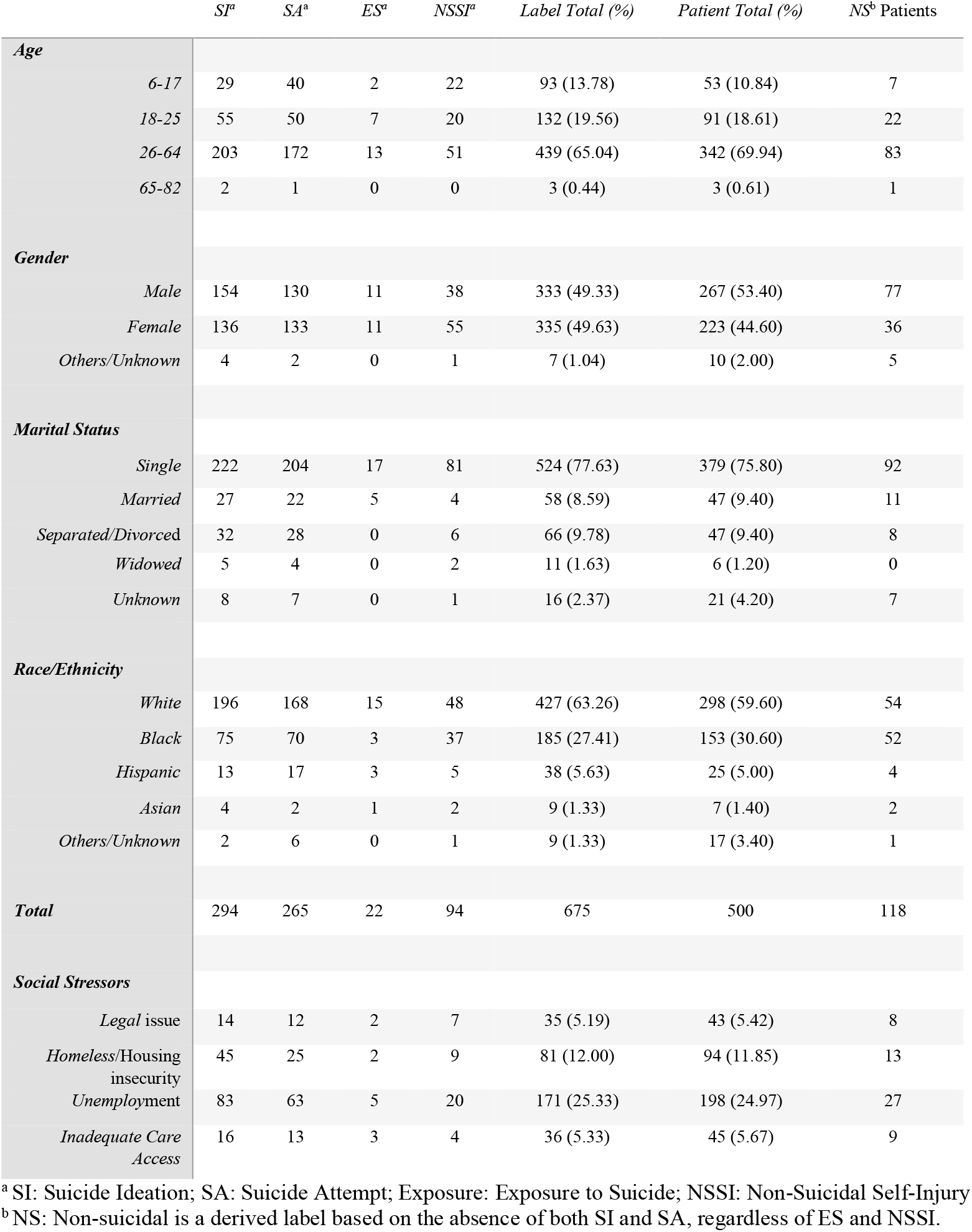
Study cohort demographics as well as social stressors by suicide-related events and factors.

### Corpus Analysis

Inter Annotator Agreement (IAA) was calculated for the first 200 notes used to develop the near-final annotation guideline with Cohen’s Kappa scores (κ) of 0.86 for SI, 0.83 for SA, 0.81 for ES, and 0.77 for NSSI. Higher IAA scores indicated better annotation consistency and/or easier annotation tasks, while lower scores resulted from challenges in the initial definitions and understanding of annotation rules. After the iterative panel discussion and knowledge refinement, the annotation guideline was finalized and applied to the entire corpus, and IAA for the 500 notes was significantly improved, with scores of 0.98 for SI, 1.0 for SA, 0.98 for ES, and 0.9 for NSSI, averaging κ = 0.95, indicating near-perfect agreement [0.81–01.00] (35). A total of 675 labels were created for the 500 IPE notes, including 294 SI, 265 SA, 22 ES, and 94 NSSI. We found a total of 118 notes labeled with NS, 103 without any of the four suicide labels and 15 with either ES or NSSI, or both. Of the 397 labeled notes, 172 (34.4%) contained one label, predominantly SI (N=96, 24.2%) or SA (N=62, 15.6%), with fewer for NSSI (N=11) and ES (N=3). Meanwhile, 225 notes (45%) featured with two or more labels, most commonly SI and SA (N=129, 25.8%). Rare combinations included ES only (N=3, 0.6%), ES and NSSI (N=2, 0.4%), SA and ES (N=2, 0.4%), SA and NSSI (N=17, 3.4%), and SI and ES (N=3, 0.6%). A total of 14 out of 16 possible label combinations were observed, missing two combinations (SA-ES-NSSI, SI-ES-NSSI). A Venn diagram (**Figure 3**) illustrated label overlaps, highlighting the coexisting nature of suicidal events and related factors.

**Figure 3.**
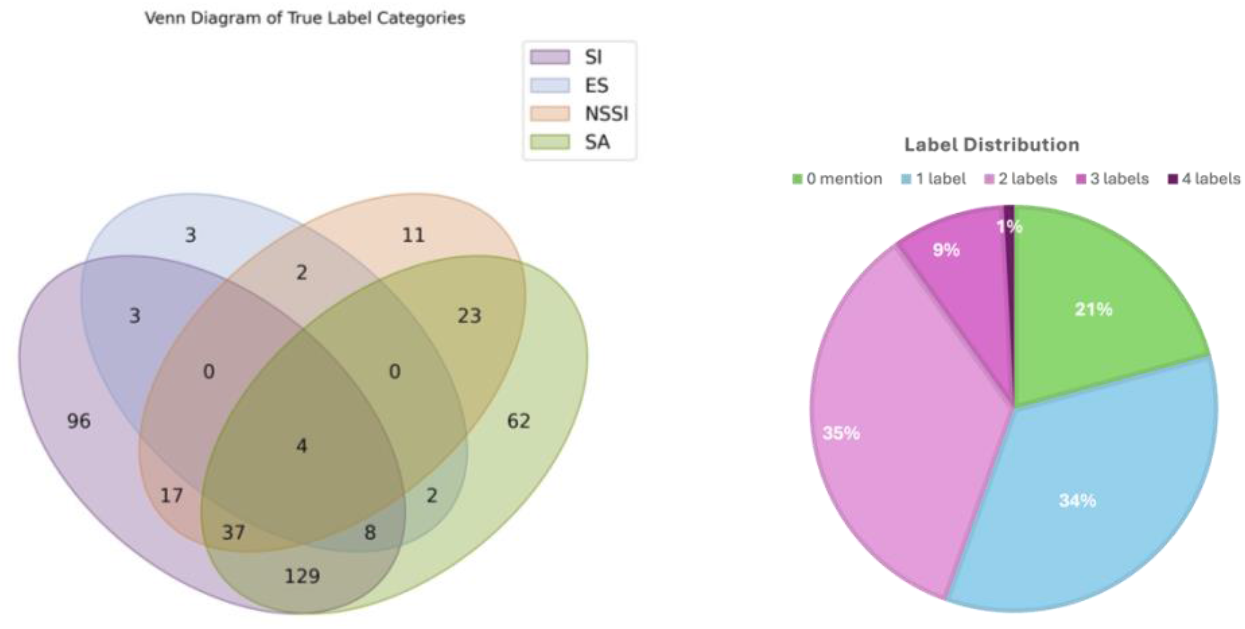
Venn diagram and pie chart for label distribution of suicide-related events and factors

### AI-powered Classification

A multi-label classifier was implemented based on BERT as the baseline model for automatic classification using the binary relevance approach, where each label is independently assigned a binary classifier. The model achieved an accuracy of 0.72±0.03 and an F1 score of 0.78±0.02 for SI, 0.73±0.04 and 0.74±0.04 for SA, and 0.81±0.01 and 0.24±0.13 for NSSI. Due to data scarcity, the model failed to predict any positive cases for ES.

## Discussion and Insights

To advance our understanding of suicide events and related factors and their documentation in the clinical notes of psychiatric safety-net hospital, we developed a clinically-sound annotation guideline and constructed an annotated corpus of 500 IPE notes. The near-perfect IAA score of 0.95 demonstrates the feasibility of our strategy based on expert panel and iterative procedure to create a high-quality annotated corpus for data analysis and modeling.

Demographic analysis revealed that a young population with a mean age of 33.42 in the study cohort. Younger individuals were more likely to exhibit NSSI (a mean age of 26.90 years), consistent with broader trends of increased self-harm among younger populations and highlighting the need for targeted interventions (36). Most patients (75.80%) were single, suggesting potential social isolation linked to suicidal tendencies and psychiatric conditions. Social determinants of health (SDoH), such as unemployment (24.2%) and homelessness (12.0%), were prevalent in the cohort, likely exacerbating psychiatric conditions and influencing suicidality. While the current study did not analyze associations or constructing predictive models, these findings emphasize the importance of future research examining the interplay between SDoH, psychological stressors, and suicidality to enhance clinical risk assessment and intervention strategies. Additionally, the co-occurrence of SI and SA (35.6%) signals a heightened suicide risk, often necessitating intensive interventions like 24-hour monitoring. Cases involving three or more labels (9%) or all four (0.8%) highlight the complex clinical scenarios faced in high-acuity psychiatric settings, complicating treatment planning and resource allocation.

During this annotation process, we identified and addressed four fundamental challenges that arose consistently via content analysis of the annotated corpus: implicit information, conflicting information, ambiguous information, and coverage of definition. These insights not only operationalized the annotation process but also have the potential to improve clinical documentation practices.

### (1) Implicit Information

Suicidal ideation and suicide-related behaviors were often not explicitly documented using direct terms like “suicide” or “kill herself”. Instead, these concepts were conveyed through more subtle, patient-reported statements such as “I want to end it all” or “plan to walk into traffic”. These indirect expressions made it difficult for traditional NLP models to capture the context of suicidality without a deeper understanding of the patient’s mental state and the nuances of the clinical expression. The guideline was developed to identify these subtleties and ensure that implicit mentions were correctly annotated, allowing for more accurate classification of suicide events and related factors. We recommend providers to explicitly use standard diagnostic terminologies to describe the observed symptoms related to suicide in clinical documentation.

### (2) Conflicting Information

Another major challenge was the frequent occurrence of conflicting information between patient-reported suicidality and expert clinical judgment. Many patients, particularly those admitted involuntarily, often minimized or denied their suicidal intentions and behaviors, likely in an effort to shorten their hospital stay or avoid more intensive intervention measures. In contrast, clinicians’ evaluations sometimes indicated objective risk, based on clinical observations or other indicators. To address this, we established a rule within the annotation guideline to prioritize expert-reported evidence in cases where there was a clear discrepancy between patient self-reports and clinical documentation. This rule helped resolve conflicts and ensured that the annotations reflected a more clinically accurate representation of the patient’s risk level. We recommend providers to include justifications or clinical rationales to support their expert judgement in the case of patients denies their suicidality.

### (3) Ambiguous Information

Furthermore, clinical notes often consist of ambiguous information where intent was unclear, particularly in differentiating between NSSI and SA. We categorized these ambiguous cases into three categories: inadequate information, mismatch between content and template, and undetermined intent. Some notes lacked sufficient detail to distinguish whether certain behaviors, such as overdoses or self-inflicted injuries, were attempts to die or simply expressions of distress without suicidal intent. Additionally, documentation templates sometimes placed NSSI behaviors under headers related to suicide attempts, further complicating the interpretation of intent. It is often the case that patients experiencing extreme psychotic episodes are unable to provide clear insights into their own intentions, including whether their actions were driven by suicidal thoughts or other factors. Their mental state may be so severely impaired that they are incapable of articulating their motives, making it difficult for providers to accurately assess whether self-injurious behaviors such as “cutting” were intended as suicide attempts or were the result of delusions, hallucinations, or other psychotic symptoms. This lack of insight poses a significant challenge in evaluating the true level of risk and complicates the documentation for accurate classification. It marks more importance to rely on clinicians’ judgements with documented justification.

### (4) Coverage of Definitions

Despite the implementation of these general rules, special cases emerged that did not neatly fit within formal definitions based on clinical guidelines and literature. One particularly challenging case was the documentation of auditory hallucinations commanding suicide. For instance, patients reported statements such as, “a demon keeps telling me to kill myself”, “I hear voices all the time that I should die”, and “God says I would not die if I jumped.” These hallucinations, often experienced by patients with severe psychiatric conditions, such as schizophrenia, present a unique form of suicidal ideation where the drive to harm oneself is not a conscious decision but a response to external, commanding voices. Such cases carry significant clinical weight and should be labeled as SI, because they indicate a higher level of risk and severity, necessitating urgent intervention.

## Limitation

This study has several limitations. First, the relatively small sample size of 500 annotated IPE notes limits the comprehensiveness of cases, with two combinations (SA-ES-NSSI, SI-ES-NSSI) absent and some combinations too infrequent for meaningful statistical analysis. Second, because the data is sourced from a single safety-net hospital, the findings may lack generalizability to broader psychiatric settings, particularly non-safety-net care. Third, the annotated corpus exhibits significant label imbalance. This reflects clinical documentation practices but poses challenges for AI models in accurately classifying less common labels like ES and NSSI. Consequently, model performance for these labels shows lower recall, potentially underestimating their true clinical significance. Fourth, we did not differentiate the temporality, severity, or frequency of the four suicide-related labels. These modifiers are critical for diagnosis and treatment planning, offering deeper insights into a patient’s risk and care needs. For instance, temporality can distinguish chronic from acute SI, while severity and frequency inform intervention urgency and recurrence risk. Clinicians are encouraged to document these descriptive modifiers to enhance patient care and support.

## Future Steps

Future studies should focus on expanding the sample size to better represent rare suicidal behaviors such as ES and NSSI, thereby improving AI model performance in detecting these less frequent but clinically significant labels. Incorporating descriptive modifiers—such as temporality, severity, frequency, and risk/protective factors, including SDoH—into annotation and analysis frameworks would provide a more nuanced understanding of suicide risk across diverse populations. Leveraging advanced language models like ChatGPT and Llama (37, 38), alongside in-context learning techniques, could enhance multi-label classification by identifying implicit mentions of suicidality and improving context-aware analysis, even with limited examples of rare cases. These models could also enable large-scale analysis of clinical notes, facilitating associative and network analyses and predictive modeling of complex factor-outcome interactions to deepen insights into suicide risk and related behaviors.

## Conclusion

In this study, we developed an annotation guideline and constructed an annotated corpus of 500 IPE notes to enhance the understanding of suicidal behaviors within a psychiatric safety-net hospital setting. By addressing critical challenges—such as label imbalance, implicit, conflicting, and ambiguous information, and the complexity of co-occurring suicide-related factors—we operationalized a framework for more accurate annotation and analysis of suicide phenotypes. Our findings highlighted the importance of standardizing clinical documentation practices to better capture subtle and nuanced expressions of suicidality, ultimately improving the quality of annotations and downstream AI applications. The high-quality annotated corpus is also de-identified and shared via data user agreement for model development and extensive validation.

While AI-powered language model demonstrated promising performance in detecting suicidal ideation and attempts, their limitations, including label imbalance and small sample size, underscore the need for larger, more diverse datasets and the adoption of advanced language models. Future work should prioritize the integration of clinically significant modifiers—such as temporality, severity, frequency, and social determinants of health (SDoH)—to refine risk assessment and enhance clinical decision-making. Additionally, leveraging state-of-the-art language models like ChatGPT and Llama, combined with context-aware techniques, could further improve the classification of implicit and complex suicide-related factors for vulnerable populations.

## Supporting information

Annotation Guideline in Supplemental material

## Data Availability

All data produced in the present study are available upon reasonable request to the authors.

https://github.com/leoliaugust1230/HCPC_IPE_labeled_suicide

## Reference

1. Klonsky ED, May AM: The three-step theory (3ST): A new theory of suicide rooted in the “ideation-to-action” framework 2015; 8:114–129

2. Sun S: Meta-analysis of Cohen’s kappa 2011; 11:145–163

3. Levis M, Westgate CL, Gui J, et al.: Natural language processing of clinical mental health notes may add predictive value to existing suicide risk models 2021; 51:1382–1391

4. Shiner B, Levis M, Dufort VM, et al.: Improvements to PTSD quality metrics with natural language processing 2022; 28:520–530

5. Posner K, Oquendo MA, Gould M, et al.: Columbia Classification Algorithm of Suicide Assessment (C-CASA): classification of suicidal events in the FDA’s pediatric suicidal risk analysis of antidepressants 2007; 164:1035–1043

6. Devlin J: Bert: Pre-training of deep bidirectional transformers for language understanding 2018;

7. Spasic I, Nenadic G: Clinical text data in machine learning: systematic review 2020; 8:e17984

8. Chim J, Tsakalidis A, Gkoumas D, et al.: Overview of the clpsych 2024 shared task: Leveraging large language models to identify evidence of suicidality risk in online posts2024, pp 177–190.

9. Portability I, Act A: Guidance regarding methods for de-identification of protected health information in accordance with the health insurance portability and accountability act (HIPAA) privacy rule 2012;

10. Poulin C, Shiner B, Thompson P, et al.: Predicting the risk of suicide by analyzing the text of clinical notes 2014; 9:e85733

11. Touvron H, Lavril T, Izacard G, et al.: Llama: Open and efficient foundation language models 2023;

12. Rifkin R, Klautau A: In defense of one-vs-all classification 2004; 5:101–141

13. Peterson C, Haileyesus T, Stone DM: Economic cost of US suicide and nonfatal self-harm 2024;

14. Fernandes AC, Dutta R, Velupillai S, et al.: Identifying suicide ideation and suicidal attempts in a psychiatric clinical research database using natural language processing 2018; 8:7426

15. Sunyang F: TRUST: clinical text retrieval and use towards scientific rigor and transparent process 2021;

16. Kariotis TC, Prictor M, Chang S, et al.: Impact of electronic health records on information practices in mental health contexts: scoping review 2022; 24:e30405

17. CDC: Facts About Suicide [Internet] Available from: https://www.cdc.gov/suicide/facts/index.html

18. Khurana D, Koli A, Khatter K, et al.: Natural language processing: state of the art, current trends and challenges 2023; 82:3713–3744

19. Association AP: Assessing and treating suicidal behaviors: A quick reference guide 2004;

20. Barak-Corren Y, Castro VM, Javitt S, et al.: Predicting suicidal behavior from longitudinal electronic health records 2017; 174:154–162

21. Schreiber, J, Culpepper L: Suicidal ideation and behavior in adults [Internet] 2024; Available from: https://www.uptodate.com/contents/suicidal-ideation-and-behavior-in-adults#references

22. Nock MK, Hwang I, Sampson N, et al.: Cross-national analysis of the associations among mental disorders and suicidal behavior: findings from the WHO World Mental Health Surveys 2009; 6:e1000123

23. Bolton JM, Gunnell D, Turecki G: Suicide risk assessment and intervention in people with mental illness 2015; 351

24. De Santis ML, Myrick H, Lamis DA, et al.: Suicide-specific safety in the inpatient psychiatric unit 2015; 36:190–199

25. Zhang Y, Zhang OR, Li R, et al.: Psychiatric stressor recognition from clinical notes to reveal association with suicide 2019; 25:1846–1862

26. Hua Y, Liu F, Yang K, et al.: Large language models in mental health care: a scoping review 2024;

27. Pandey B, Pandey DK, Mishra BP, et al.: A comprehensive survey of deep learning in the field of medical imaging and medical natural language processing: Challenges and research directions 2022; 34:5083–5099

28. Windfuhr K, Kapur N: Suicide and mental illness: a clinical review of 15 years findings from the UK National Confidential Inquiry into Suicide 2011; 100:101–121

29. Adekkanattu P, Furmanchuk A, Wu Y, et al.: Deep learning for identifying personal and family history of suicidal thoughts and behaviors from EHRs 2024; 7:260 30.

30. Spasic I, Nenadic G: Clinical text data in machine learning: systematic review 2020; 8:e17984

31. Coppersmith G, Leary R, Crutchley P, et al.: Natural language processing of social media as screening for suicide risk 2018; 10:1178222618792860

32. Swain RS, Taylor LG, Braver ER, et al.: A systematic review of validated suicide outcome classification in observational studies 2019; 48:1636–1649

33. Wu S, Roberts K, Datta S, et al.: Deep learning in clinical natural language processing: a methodical review 2020; 27:457–470

34. Castelpietra G, Knudsen AKS, Agardh EE, et al.: The burden of mental disorders, substance use disorders and self-harm among young people in Europe, 1990–2019: Findings from the Global Burden of Disease Study 2019 2022; 16

35. Thatoi P, Choudhary R, Shiwlani A, et al.: Natural Language Processing (NLP) in the Extraction of Clinical Information from Electronic Health Records (EHRs) for Cancer Prognosis 2023; 10:2676–2694

36. Andriessen K, Rahman B, Draper B, et al.: Prevalence of exposure to suicide: A meta-analysis of population-based studies 2017; 88:113–120

37. Ray PP: ChatGPT: A comprehensive review on background, applications, key challenges, bias, ethics, limitations and future scope 2023; 3:121–154

38. Cerel J, Maple M, van de Venne J, et al.: Exposure to suicide in the community: Prevalence and correlates in one US state 2016; 131:100–107

