## Supplementary material for "INSIGHTFUL: Insight Generation through Clinical Annotation, Analysis, and Modeling of Suicide-Related Factors towards Understanding and Lifesaving": Annotation Guideline in Supplemental material

Supplement

**Data Analysis**

Social Stressors were extracted as diagnosis codes in the structured EHR. Stressor categories captured aggregates of related diagnosis codes. “Legal Issue” covers ‘legal problems’, ‘litigation’, ‘incarceration’, ‘on parole’; “Homeless/Housing Insecurity” covers ‘inadequate housing’, ‘housing issue’, ‘homelessness’, ‘homeless’; “Unemployment” includes ‘job stress’, ‘unemployment’, ‘severe financial problem’, and ‘financial stressors.’; and “Inadequate Care Access” covers ‘inadequate health care access,’ and ‘access to health care.’.

**Model training specification**

In this study, we implemented an AI-powered multi-label classification approach to identify coexisting suicidal events and related factors at the document level. We utilized Bidirectional Encoder Representations from Transformers (BERT) models with a supervised learning strategy. Specifically, we employed a binary relevance method, also known as One-vs-All, where each suicidal event was treated as a separate binary classification task. This approach converts a multi-label dataset into multiple single-label binary datasets.

We fine-tuned each model on a training set of 400 IPE notes and evaluated their performance on a test set of 100 notes. To enhance the reliability of our results and reduce sampling bias, we used Repeated Stratified K-Fold cross-validation. The training and evaluation were performed using 5-fold cross-validation with 3 repetitions. The training process employed the following hyperparameters: a learning rate of 1e^5^, a batch size of 4, and 5 training epochs, with a weight decay of 0.01 using the AdamW optimizer. The maximum token input length was set to 512 tokens, with longer texts truncated as needed. Notably, 431 (86.2%) of the notes had less than 512 tokens.

**Annotation Guideline for Identifying Suicide-related Events From Clinical Notes**

Revision 3

08/13/2024

**Background**

Unstructured EHR clinical notes contain a wealth of information on the suicidality of persons with psychiatric conditions. However, it is time-consuming and costly to manually extract this information from clinical records for large patient populations. In this study, we will mine a safety-net psychiatric hospital EHRs using NLP techniques to identify the presence of suicide-related factors and events among persons with psychiatric admissions. Our goal is to develop a novel clinical corpus and evaluate NLP algorithms to identify characteristics of patients with various suicidalities from unstructured clinical notes at the Harris County Psychiatric Hospital (HCPC). This annotation guideline was derived from peer-reviewed meta-analysis^1^, clinical guideline ([Wolter Kluwer’s UpToDate](https://www.uptodate.com/contents/suicidal-ideation-and-behavior-in-adults)), and a standardized suicidal rating system ([Columbia Classification Algorithm of Suicide Assessment](https://www.ncbi.nlm.nih.gov/pmc/articles/PMC3804920/)). This guideline was reviewed and revised by four board-certified psychiatrists.

**Annotation Task**

Please use this document for annotation to make sure we are consistently collecting the same information in the same way. Perform document-level, multi-label classification to identify four suicide-related events and factors in clinical notes. Each document should be annotated with the presence (1) or absence (0) of the four categories related to suicide mentions. Annotation will only take place within the secure data storage environment (UTHealth Google Drive) using Google Sheets.

Table 1: Conceptual annotation sample

| **Note** | **SI** | **SA** | **ES** | **NSSI** |
| --- | --- | --- | --- | --- |
| Text 1 | 0 | 1 | 1 | 1 |
| Text 2 | 0 | 0 | 1 | 0 |
| Text ... | 1 | 1 | 0 | 1 |

**Labels**

1. **Suicide Ideation (SI)**
2. **Suicide Attempt (SA)**
3. **Exposure to Suicide (ES)**
4. **Non-Suicidal Self-Injury (NSSI)**

| **Case** | **Sample de-ided Text** | **SI** | **SA** | **ES** | **NSSI** |
| --- | --- | --- | --- | --- | --- |
| 1 | [Patient] bought in from [Hospital] for OD ing on Seroquel (12x 50mg), Prozac (35 x 10mg), Tylenol (39x 500mg) behind closed doors in their house. 1st SA.  Of note GCS was 8 on [Date] and changed to 12 in the ER. Got into argument with mother. Self mutilation multiple times (to relieve inner pain) Sexual abuse by mother's BF. Used to get good grades but now believes they are going down.  Patient endorses depressed mood; poor sleep; reports feelings of hopelessness, helplessness, worthlessness; SI. Mother does not listen to me much and feel she ignores me. "If she won't listen to me maybe she will listen to me dying."  Not close to anybody as I choose not to. Past history of Depression (diagnosed in [Date]). Prior psych hospitalization: [Duration] at [Hospital] for SI in [Date]. Currently on Seroquel (50mg qPM) and Prozac (30mg qAM). Stopped Seroquel a few days ago. F/u with Dr. [Name] and Dr. [Name] Denies any past or current recreational or IV drug abuse | 1 | 1 | 0 | 1 |
| 2 | [Age] y/o individual was transferred involuntarily from [Facility] where they were taken after voicing thoughts indicating suicidal ideation (SI). Per intake documents, the patient had been consuming alcohol (ETOH) prior to expressing to their spouse and child that "I don't want to be here". It was also reported that the suicide of the patient's brother in [Date] was contributing to their depression. The patient denied any history of prior psychiatric treatment. Xanax had been prescribed for "Anxiety" following the brother's death. Tobacco use: 2 packs per day (ppd). ETOH: "only socially" (6-pack every weekend)/ last consumed on [Date] History of 2 DUIs [Time Frame] ago; Rehabilitation x1; no history of withdrawals. Denies any other past or current recreational or IV drug abuse. | 1 | 0 | 1 | 0 |

Table 2. Sample annotation with deidentified real cases

**General Rules**

**Inclusive Criteria**

- **Literal Interpretation:** Assign labels based on **the explicit content** of suicide-related events in the clinical notes. Avoid making inferences beyond what is written.
- **Temporal Agnostic:** Annotate any mentions of both current and lifetime suicidal events.
- **Severity Agnostic:** Annotate any mentions of suicidal events regardless of severity.
- **Frequency Agnostic:** Annotate any mentions of suicidal events regardless of frequency.
- **Prioritization of Evidence:** If a clinical note showed multiple evidence, including physician's observation, family/friends' reports, and/or patient's self-report, the annotators will make a decision in the following order of evidence:

**Physician's Observations/Judgments ……………….**

1^st^

2^nd^

3^rd^

**Reports from Patient's Family/Friends……………**

**Patient's Self-Report …………………………….**

*Example: “She minimized the events surrounding her suicide attempt, saying that she was just trying to go to sleep”.*

Explanation: In this case, **SA** should be annotated because the physician deemed the event as a **suicide attempt** while the patient minimized the intention.

See more examples in page 8.

**Exclusive Criteria**

- Other risk factors related to suicide, such as being hopeless, having a history of depression, or going through a bad breakup.
- The patient denies any suicidal events **AND** there are no physician-reported indications of suicide. Annotate all labels with “0”.

*Examples: "Denies SI.", "He denies S/HI."*

**Label Definitions and Examples**

**1. Suicide Ideation (SI)**

- **Formal Definition (UpToDate):** Thoughts about killing oneself; these thoughts may include a plan.
  - **Suicide threat (UpToDate):** Thoughts of engaging in self-injurious behavior that are verbalized and intended to lead others to think that one wants to die, despite no intention of dying.
- **Working Definition:** Any mention by the patient or physician of thoughts about killing oneself. This includes suicide threats, written suicide notes, and social media posts as well as.
- **Special Note:** In the case when SI is not from the conscious mind, but due to psychiatric conditions such as auditory hallucination commanding patients to kill self, include these instances as positive SI cases.

Table 3. Suicide Ideation (SI)

| **Construct** | **Description** | **Examples** |
| --- | --- | --- |
| Suicide Ideation | thinking about, considering, or planning suicide | - "I have been thinking about killing myself." - "I want to end my life." - "Recurrent thoughts of death." - “he had a plan to slit his wrist with a blade” |
| Suicide Threats | A verbal statement said to other people about one’s suicidal tendency | - "Pt voicing suicide threat. He threatened to jump off an overpass to kill himself." - “hx presents with reports of suicidal threats, AH and noncompliance.” |
| Social Media Post | A digital statement indicating suicidal tendencies | “He says he did not intend to show anyone the suicide note, posted on Facebook that he was planning to finish writing this note” |
| Suicide Note | A written or printed statement indicating suicidal tendencies | “He denies any desire to end life now but admits he wrote a suicide note, says he just wanted to write the note.” |
| Auditory Hallucination | Voice in mind without conscious control | - "His auditory hallucinations are derogatory in nature, asking him to kill himself." - “Per records patient is hearing voices telling him to hurt himself, he voiced suicidal thoughts with plan.” - “He endorsed seeing shadows and hearing voices asking to kill him” |

**2. Suicide Attempt (SA)**

- **Formal Definition (C-CASA):** A potentially self-injurious behavior, associated with at least some intent to die, as a result of the act. Evidence that the individual intended to kill him/ herself, at least to some degree, can be explicit or inferred from the behavior or circumstance. A suicide attempt may or may not result in actual injury.
- **Working Definition (UpToDate):** Any mention by the patient or physician of self-injurious behavior intended to die but did not result in fatality. (emphasis on taking actions)
- **Special Note:** Overdose (OD), when not described as a medical or accidental event, is a positive SA case.

Table 4. Suicide Attempt (SA)

| **Construct** | **Description** | **Examples** |
| --- | --- | --- |
| Suicide Attempt   - Firearm - Poisoning - Hanging - Cutting - Jump - Others | Actions taken toward killing oneself | - “0 previous admissions, per involuntary basis due to suicide attempt” - “stated pt had been violent in the past and had also attempted suicide in the past” - "Trying to swallow paper clips and cut himself." - “he had a suicide attempt a year ago when he tried to slit his throat” - “He pickedup a gun in his closet, said goodbye to his children and cousin while his wife called 911.” - “patient was stabilized after a self-inflicted gunshot wound” - “HX OF SUICIDE ATTEMPTS: last time couple of years ago by slit wrist, hang self, jumped in front of traffic” |
| Overdose | Intentionally ingest excessive amount of substance to kill oneself | - "Patient admitted due to overdose, attempting suicide." - "Several previous suicide attempts by OD, hanging, pouring gasoline on self." - "Patient came in intoxicated following drug overdose. Overdose was intentional." - "She ingested 7-8 Ibuprofen in a possible suicide attempt." - “patient had taken 148 pills of asprin in a attempt to kill himself” |
| Non-suicidal Overdose **(exclude)** | a medical or accidental use of excessive amount of substance | - “Denies past SA although unintentionally/accidentally overdosed in '81 on his sleeping meds” - “has recently accidentally overdosed on Trileptal and Seroquel in past week” |

**3. Exposure to Suicide (ES)**

- **Formal Definition (Meta-Analysis**^1^**)**: Having experienced a suicide among family or friends, or having personally known someone who has died through suicide.
- **Working Definition:** Any mention of suicide-related events (SI, SA, and suicide death) experienced by individuals other than the patient, such as family members or friends.
- **Special note:** Deaths that are not indicated as suicide should be excluded.

Table 5. Exposure to Suicide (ES)

| **Construct** | **Description** | **Examples** |
| --- | --- | --- |
| Exposure to Suicide death | Family members, friends, or colleagues of a person who died by suicide | - "Family history of suicide." - "He reportedly witnessed suicide of his brother who shot himself in front of him while playing Russian roulette making suicidal duel." - “He says that his aunt committed suicide in April 2007 and his life has never been the same” - “blames himself for father committing suicide” - “Pt said that he started feeling suicidal after his friend shot himself” |
| Exposure to Suicidal tendency | Family members, friends, or colleagues of a person who experience suicidal tendencies, such as SI and SA. | - “This brother of hers tried to kill himself 3 years ago because of the family conflicts they have.” - “first depressive episode was at age 15 yo when she saw her father trying to cut his neck” |
| Exposure to non-suicide death **(exclude)** | Family members, friends, or colleagues of a person who died by other causes instead of suicide | - “Her mother died when pt. was 5 yr. of age.” - “grandfather died 2 days ago” - “patient said he had needed "psychiatric treatment" since his cousin was murdered.” |

**4. Non-Suicidal Self-Injury (NSSI)**

- **Formal Definition (UpToDate)**: Self-injurious behavior characterized by the deliberate destruction of body tissue in the absence of any intent to die and for purposes that are not socially sanctioned.
  - **Non-suicidal self-injurious thoughts**: Thoughts of engaging in NSSI.
- **Working Definition:** Any mention of deliberate self-inflicted harm to body tissue without intent to die and not socially sanctioned. This includes non-suicidal self-injurious thoughts as well.
- **Special Note:**
  - NSSI due to impulsivity should also be included as a positive NSSI case.
  - Non-suicidal self-injurious thoughts should also be included as a positive NSSI case.
  - Exclude broader types of self-harming or unhealth behaviors, such as engaging in emotionally abusive relationships, substance abuse, or gaming addiction.
  - Exclude medical or accidental injury caused by self, such as accident overdose,

Table 6. Non-Suicide Self-Injury (NSSI)

| **Construct** | **Description** | **Examples** |
| --- | --- | --- |
| Non-Suicidal Self-Injury   - Cutting/slitting - Hitting/banging - Scratching | deliberately injuring oneself without suicidal intent | - "Cutting himself on his leg once before to relieve tension." - “She has never attempted suicide but said she has made superficial cut to her wrist in the past but said this was not to kill herself" - She used to cut herself to release some of her emotional pain." - "He cut on his forearm with razor several times." - "Hx Self-mutilation." - “hitting her self for agitation” - “he had commented that he would hurt himself” - “has a h/o self-injurious behavior (banging head against wall "when I am angry") but denies any h/o cutting” - “starts scratching his forearms, he also reports that he pulls at his skin.” |
| General unhealthy behaviors **(exclude)** | Self-aware unhealthy behaviors without intention to self-harm | - "Pt. has hx of opiate and benzo abuse." - “*Pt smokes 10 cigs/day” - “Patient with a past psychiatric history of Cocaine abuse” - “has h/o polysubstance abuse” - “He has not slept in 4 days and has started to loose his mind” |

**Handling Ambiguous and Challenging Cases**

- **Unclear Intentions:** If the intention behind self-injury behavior is ambiguous, annotate both **SA** and **NSSI** based on the explicit information provided.

Table 7. Ambiguous Cases

| **Ambiguity** | **Examples** | **Annotation Decision** |
| --- | --- | --- |
| Not enough information | “*History of suicide attempts/self-injury: few months ago” | SA, NSSI |
| NSSI information in SA template | “Suicide attempts: slit wrists but knew slitting wrists wouldn’t kill, says it was like a “cry for help”.” | SA, NSSI |
| Undetermined intent | “Patient has cut/self-injured 3 times with unclear want to die.” | SA, NSSI |

**Conflicting Information:** When conflicting details present, follow the general priority rules defined on page 2, prioritize physician assessments and Previous medical record as the highest level of evidence.

Table 7. Conflicting Cases

| **Priority type** | **Examples** | **Annotation Decision** |
| --- | --- | --- |
| Physician’s judgement | “she endorsed SI with plans to hang herself with a rope … Denies SI and depressive symptoms…” | SI |
| Family member report | “patient's mother called because patient threatened to kill herself and then sent suicidal texts, patient denies that she threatened to kill her mother or herself” | SI |
| Previous medical record | “Pt presents after ingesting 12 Tylenol 500mg pills in a one hour time frame. The pt states "I wasn't trying to kill myself. I just had a massive headache." … Records from the hospital show the pt admitted to depression and intentionally overdosing.” | SA |

**Reference**

1. Andriessen K, Rahman B, Draper B, Dudley M, Mitchell PB. Prevalence of exposure to suicide: A meta-analysis of population-based studies. *J Psychiatr Res*. 2017;88:113–120.
